# Perceived vulnerability to COVID-19 infection from event attendance: Results from Louisiana, USA, two weeks preceding the national emergency declaration

**DOI:** 10.1101/2020.04.01.20049742

**Authors:** Ran Li, Bingcheng Yang, Jerrod Penn, Bailey Houghtaling, Juan Chen, Witoon Prinyawiwatkul, Brian E. Roe, Danyi Qi

**Affiliations:** Department of Agricultural Economics and Agribusiness, Louisiana State University and LSU AgCenter, Baton Rouge, Louisiana, United States of America; School of Nutrition and Food Sciences, Louisiana State University and LSU AgCenter, Baton Rouge, Louisiana, United States of America; Department of Agricultural, Environmental and Development Economics, Ohio State University, Columbus, Ohio, United States of America; School of Business, Sun Yat-sen University, Guangzhou, China; Department of Human Resource Management, College of Humanities, Sichuan Agricultural University, Yaan, Sichuan, China

## Abstract

In response to the mounting threat of COVID-19, we added questions to an ongoing food preference study held at Louisiana State University from March 3-12 of 2020. We asked 356 participants: (1) In your opinion, how likely is it that the spread of COVID-19 (the coronavirus) will cause a public health crisis in the United States? (2) How concerned are you that you will contract COVID-19 by attending events on campus? Participants’ estimates of an impending national health crisis increased significantly during the study’s second week (March 9-12) while concern about personally contracting COVID-19 from attending campus events increased only marginally during the study’s final days. We find those expressing a higher likelihood of an impending national crisis were more concerned about contracting COVID-19 by attending campus events, suggesting a possible transmission from perceptions of national-level events to perceived personal vulnerability via local exposure. However, about 30% of participants perceived that COVID-19 would likely cause a public health crisis yet did not express concern about contracting COVID-19 from event attendance. These participants were significantly more likely to be younger students who agreed to participate in response to recruitment using same day flyer distribution. Women expressed a higher likelihood of an emerging national health crisis, although they were not more concerned than men that attending campus events would result in virus contraction. Other groups (e.g., white, students younger than 25, highest income group) displayed similar concern about a national-level crisis, yet were significantly less concerned about contracting COVID-19 from attending campus events than others. Also, participants randomly assigned to information emphasizing the national impacts of food waste expressed significantly greater concern of contracting COVID-19 by attending campus events. These results provide some initial insight about how people perceived national and personal risks in the early stages of the COVID-19 crisis in Louisiana.

## Introduction

Individual perceptions of personal and national threats posed by the transmission of SARS CoV-2 and its sequalae (COVID-19) have undoubtedly shaped the public’s initial response to and ultimately the speed and geographical diffusion of the most disruptive public health crises in the past century[1]. Cowper[2], responding to unfolding events in the United Kingdom, notes that public reaction to national level communications will critically impact how the pandemic unfolds. Bagnoli, Lio and Sguanci [3] showed that individual perception of infection risk is a critical parameter for predicting the spread of epidemics and argued for inclusion of such perceptions in epidemiological models. Zhang, Deng and Zhang [4] analyzed minor differences in COVID response times across Chinese provinces during early 2020 and found that a single-day delay in provincial response significantly increased the newly confirmed case rate by 2.2% which translates to on average of 497 more confirmed cases per 10,000 population per square kilometer. Rapid response ultimately relies upon broad-based compliance by the population, which stems from the perceived risk of the evolving phenomenon from each individual.

Previous research has documented several empirical regularities in human response during epidemics. For example, Moran and Del Valle’s [5] meta-analysis revealed that women were about 50% more likely to adopt non-pharmaceutical protective responses (e.g., mask wearing, hand washing) during respiratory epidemics. de Zwart et al. [6] studied results from surveys during the Avian Influenza (AI) epidemic and found Dutch participants were more likely to undertake preventative actions among those who were older, had less formal education, had obtained a flu vaccine, perceived higher severity of AI, perceive greater vulnerability to AI, and thought more about AI. During the H1N1 influenza epidemic in Korea, Park et al. [7] found female students reported higher perceptions of illness severity and of personal susceptibility to infection than men.

However, little is currently known about how individuals assess the national and personal risks associated with the COVID-19 pandemic during critical communications windows. To our knowledge, the only study that examines perceptions of the COVID-19 threat come from surveys in China documenting demographic correlates of psychological impacts caused by the COVID crisis [8]. These authors find that women, students, those reporting specific physical symptoms and those with unfavorable self-rated health reported significantly greater psychological impacts of COVID-19.

In this article, we share results from responses gathered during a study conducted on the campus of Louisiana State University from March 3 to March 12, 2020, a period closely preceding the closure of in-person classes and events on its Baton Rouge campus. In response to the mounting threat of COVID-19 in the United States, we added two exit questions to an ongoing in-person food preference study being held on campus. We asked 356 participants: (1) In your opinion, how likely is it that the spread of COVID-19 (the coronavirus) will cause a public health crisis in the United States? and (2) How concerned are you that you will contract COVID-19 by attending events on campus? We juxtapose the evolution of responses to these questions with official government pronouncements concerning COVID-19. We also use regression analyses and classification tree analyses to explore associations between responses to these questions and participant demographic characteristics as well as experimental treatments randomly assigned to participants as part of the ongoing study.

## Methods

### Measurement

Data were collected from participants in an ongoing study focused on understanding consumer food choice and consumption behavior during midday meals (11:00 AM – 2:00 PM). Students, staff and faculty of Louisiana State University (LSU) were recruited to participate in a study held at the campus’s Food Sensory Services Lab in which they would be offered a choice among several commercially prepared lunch options. They were provided a fixed budget for lunch and kept any unspent budget as cash compensation. After providing informed consent, participants moved to isolated, individual kiosks with a computer to answer an online survey in which information treatments were randomly assigned and subjects chose among a series of competing lunch options. One of the participant’s preferred lunch options was delivered by staff to the kiosk. Upon completing the meal, staff removed the food tray and the participant completed an online exit survey via the kiosk computer that focused on satisfaction with the provided meal and personal information.

The food preference study initially began on February 17, 2020. In late February, as concerns about the spread of COVID-19 in the United States increased, we added questions to the exit survey (S1 Appendix) to understand if these events were altering the profile of individuals who chose to participate in the study. Two questions were added: (1) In your opinion, how likely is it that the spread of COVID-19 (the coronavirus) will cause a public health crisis in the United States? (*National Likelihood*); and (2) How concerned are you that you will contract COVID-19 by attending events on campus (*Local Vulnerability*)? Participants answered these questions in sessions from March 3 to March 12, 2020, the final day of the study. LSU continued all in-person classes and food service operations through March 13, 2020, and no official announcements were made regarding the cancellations of any on-campus activities before the end of our last study session (2: 00 PM March 12^th^) [9]. At 4:00 PM on March 12^th^, 2020, LSU’s official communications regarding COVID-19 first mentioned the cancellation of on-campus classes starting from the week of March 16^th^ [10], and then announced the cancellation of non-class activities involving 30 people or more immediately at 11:30 AM on March 13, 2020 [11]. For reference, a national emergency was declared in response to COVID-19 the afternoon of March 13, 2020 [12].

### Sampling

The sample includes the 356 participants enrolled from March 3 through March 12, 2020. Individuals were recruited via pre-existing email recruitment lists, flyers circulated on campus, advertising announcements on classes, and advertisements in university locations. Inclusion criteria included age 18 years or older with no dietary restrictions to beef products.

### Analysis

Results are analyzed in Stata (version 16). The focal variables relating to COVID-19 were captured using a 5-point Likert scale. When more convenient for exposition or analysis, these responses are simplified into binary variables (very or moderately likely/concerned = 1; all other responses = 0). We also define the variable *National, Not Local* to equal one when participants think a national crisis is very or moderately likely but are neither very nor moderately concerned about contracting the virus by attending campus events. Personal characteristics included in the analyses include sex, age, student status (=1 if enrolled in University classes, =0 otherwise), household income, race, health insurance status, recycling frequency, experience with food composting, previous knowledge of food waste as an issue, whether they are trying to eat healthier, and whether they attended the session in response to in-person flyer distribution on the experiment date (as opposed to alternative recruitment such as emails or class announcements). Randomly assigned between-subjects experimental elements included in the analyses include whether participants received information about food waste (vs. screen time, *Food Waste Info*); received information about improving nutrition (vs. financial literacy, *Nutrition Info*); received meals with more vegetables (vs. fewer, *Vegetable Group*); received meals on a large plate (vs. smaller, *Large Plate*); received meals on a compostable plate (vs. plastic, *Compostable Plate*); and received menus where the vegetable was listed at the top in the description of the offering (vs. lower, *Veg Top of Menu*). More detail and context concerning the experimental elements are included in the Supporting Information (S2 appendix). The day of the study (e.g., March 3, March 4, etc.) is also controlled in all analyses. Descriptive statistics for the variables appear in Table 1.

**Table 1.** Sample Descriptive Statistics.

| VARIABLES | Mean or % |
| --- | --- |
| Dependent Variables: |  |
| National Likelihood (Likert scale, 1-5) | 3.89 |
| National Likelihood (converted to binary) | 0.74 |
| Local Vulnerability (Likert scale, 1-5) | 3.22 |
| Local Vulnerability (converted to binary) | 0.51 |
| National, not Local (converted to binary) | 0.29 |
| Female | 58.4% |
| Age × Education: |  |
| 18-24 × Non-student | 24.3% |
| 18-24 × Student | 57.8% |
| 25+ × Non-student | 7.4% |
| 25+ × Student | 10.5% |
| Household income per year: |  |
| Less than \$15,000 | 17.3% |
| \$15,000-\$49,999 | 26.9% |
| \$50,000 - \$99,999 | 14.7% |
| \$100,000 and above | 14.5% |
| Prefer not to answer | 26.6% |
| Race/Ethnicity |  |
| White | 52.7% |
| Black | 21.5% |
| Other | 25.8% |
| Hispanic or Latino | 8.8% |
| Asian | 12.5% |
| All other responses | 4.5% |
| Health insurance = yes | 88.1% |
| Recycle (sometime, about ½ the time, most of the time, or whenever possible) | 89.2% |
| Ever lived in a household that composts food | 28.6% |
| Heard about food waste | 46.7% |
| Attempt to eat a healthy diet (agree) | 80.2% |
| In-person recruitment | 40.2% |
| Randomly Assigned Experimental Elements |  |
| Food Waste Info | 50% |
| Nutrition Info | 50% |
| Vegetable Group | 32% |
| Large Plate | 63% |
| Compostable Plate | 49% |
| Veg Top of Menu | 45% |

| Study Date |  |
| --- | --- |
| Mar 3<sup>rd</sup> | 17.0% |
| Mar 4<sup>th</sup> | 13.0% |
| Mar 5<sup>th</sup> | 10.2% |
| Mar 9<sup>th</sup> | 14.2% |
| Mar 10<sup>th</sup> | 12.8% |
| Mar 11<sup>th</sup> | 17.0% |
| Mar 12<sup>th</sup> | 15.9% |
| # of Observations | 353 |

To model the Likert-scale response to the two COVID-19 perception questions, an ordered logit regression model is estimated with the aforementioned explanatory variables. The *National, not Local* response pattern model is estimated with a logit regression. Classification tree analysis is conducted for the binary version of the *Local Vulnerability*, where the Gini improvement measure is used as the splitting criteria [13]. Three participants are omitted from several analyses because of item non-response on at least one variable, leaving an effective sample size of 353. Statistical significance was set at the 5% level with results at the 10% level deemed marginally significant.

*Notes*: See supporting information for question wording and response options and for experimental element descriptions (S1 appendix and S2 appendix).

### Ethics Statement

This study was approved by the Louisiana State University AgCenter and Ohio State University Institutional Review Boards. All participants signed informed consent forms after being briefed on the study and having any questions answered by research staff. The two questions added on March 3, 2020, were granted post-hoc IRB approval.

## Results

Figure 1 depicts the number of presumptive positive COVID-19 cases reported both nationally (line graph, right axis) and in Louisiana (bar graph, left axis) for the study period, while Figure 2 traces the daily averages among study participants for the two COVID-19 questions, while highlighting key events in the evolution of COVID-19 timeline for Louisiana. Specifically, the gray bar depicts the percent who responded that COVID-19 was likely (moderately or very) to cause a national public health crisis while the black bars capture the percent that were concerned (moderately or very) that attendance at campus events would cause them to contract COVID-19.

**Fig 1.**
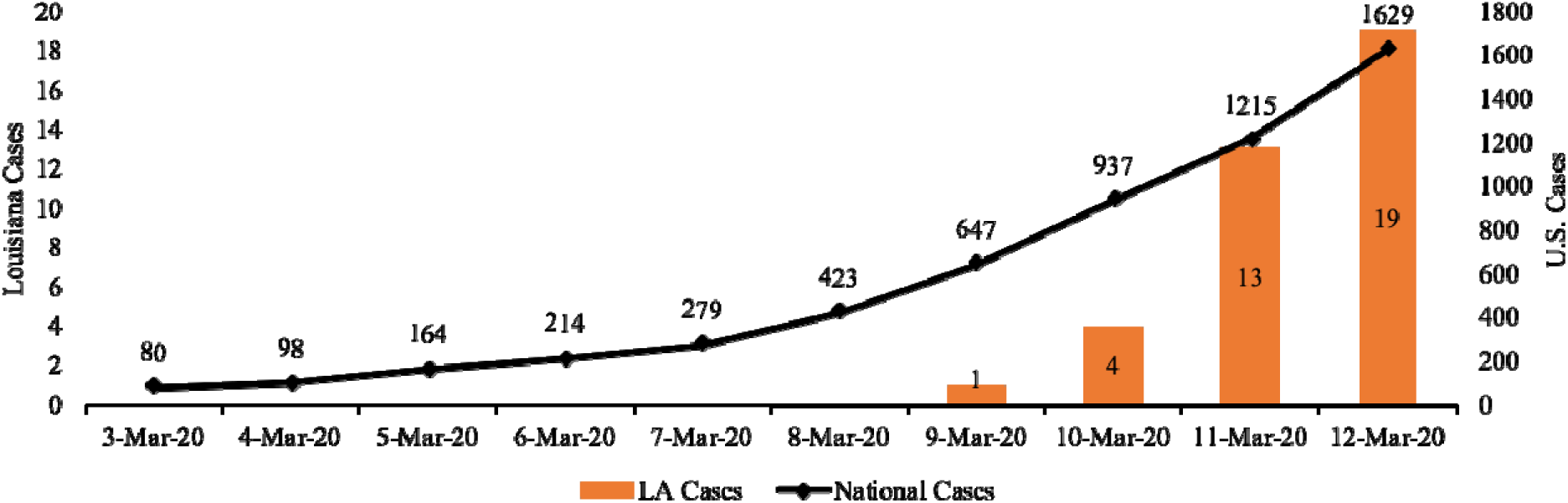
Confirmed Presumptive Positive COVID-19 Cases Reported During Study Period: Louisiana and Nationally. Data source: Centers for Disease Control and Prevention (CDC)[14], State of Louisiana: Office of the Governor[15-18]

**Fig 2.**
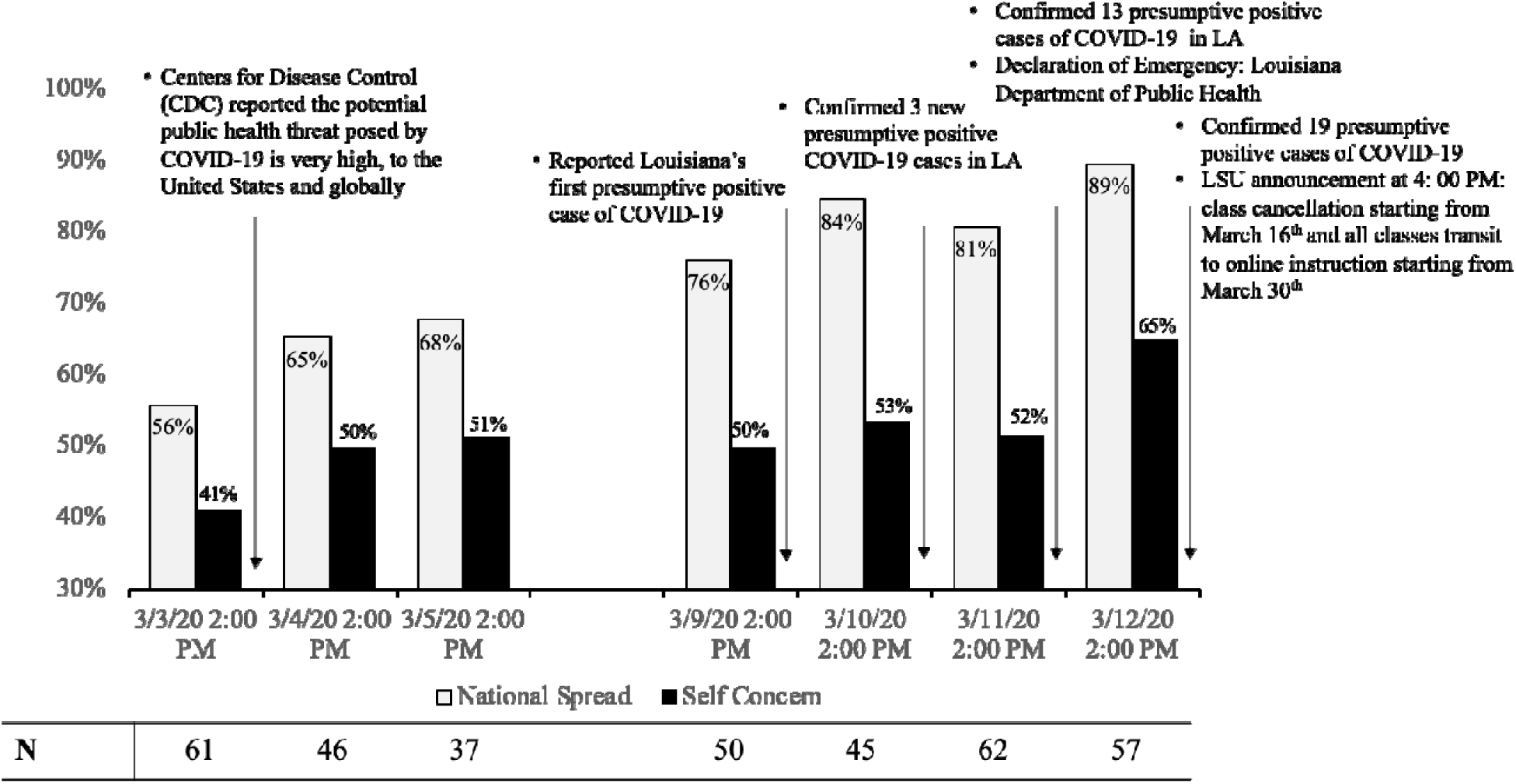
Responses to COVID-19 Questions by Study Day. *Note*: Gray bars depict percent who respond moderately or very likely that COVID-19 will cause a national public health crisis and black bars are the percent who respond moderately or very concerned that about contracting COVID-19 from attending campus events. Public announcements occurred after daily study hours, which ended by 2 PM central. Information sources: Centers for Disease Control and Prevention (CDC)[19], State of Louisiana: Office of the Governor[15-18], Louisiana Department of Health, LSU Coronavirus Updates & Infomration[10-11].

Figure 1 shows the national case count went from less than 100 on the first day of the study (March 3) to more than 1600 cases by the last day of the study (black line). Figure 2 juxtaposes the daily responses to the COVID-19 questions with key events in the national and Louisiana crisis timeline. No cases were identified and reported in Louisiana until the second week of the study (bars, Fig 1) and LSU communications stated that no cases had been identified on campus [9]. However, a lack of testing in the United States and in Louisiana likely underrepresented the prevalence of COVID-19 at the time [19].

Figure 3 shows the mean of *National Likelihood* (binary version), *Local Vulnerability* (binary version), *and National, not Local* over the experimental timeframe. *National Likelihood* increased steadily through the study period, though even on the final day of the study, more than 10% of participants did not agree that a national crisis was likely. *National Likelihood* was statistically greater than the first day of the study from March 9 to March 12, i.e., the entire second week of the study period. Statistical significance was determined from the regression model (Table 2) which controls for personal characteristics and experimental conditions randomly assigned as part of the study.

**Table 2.**
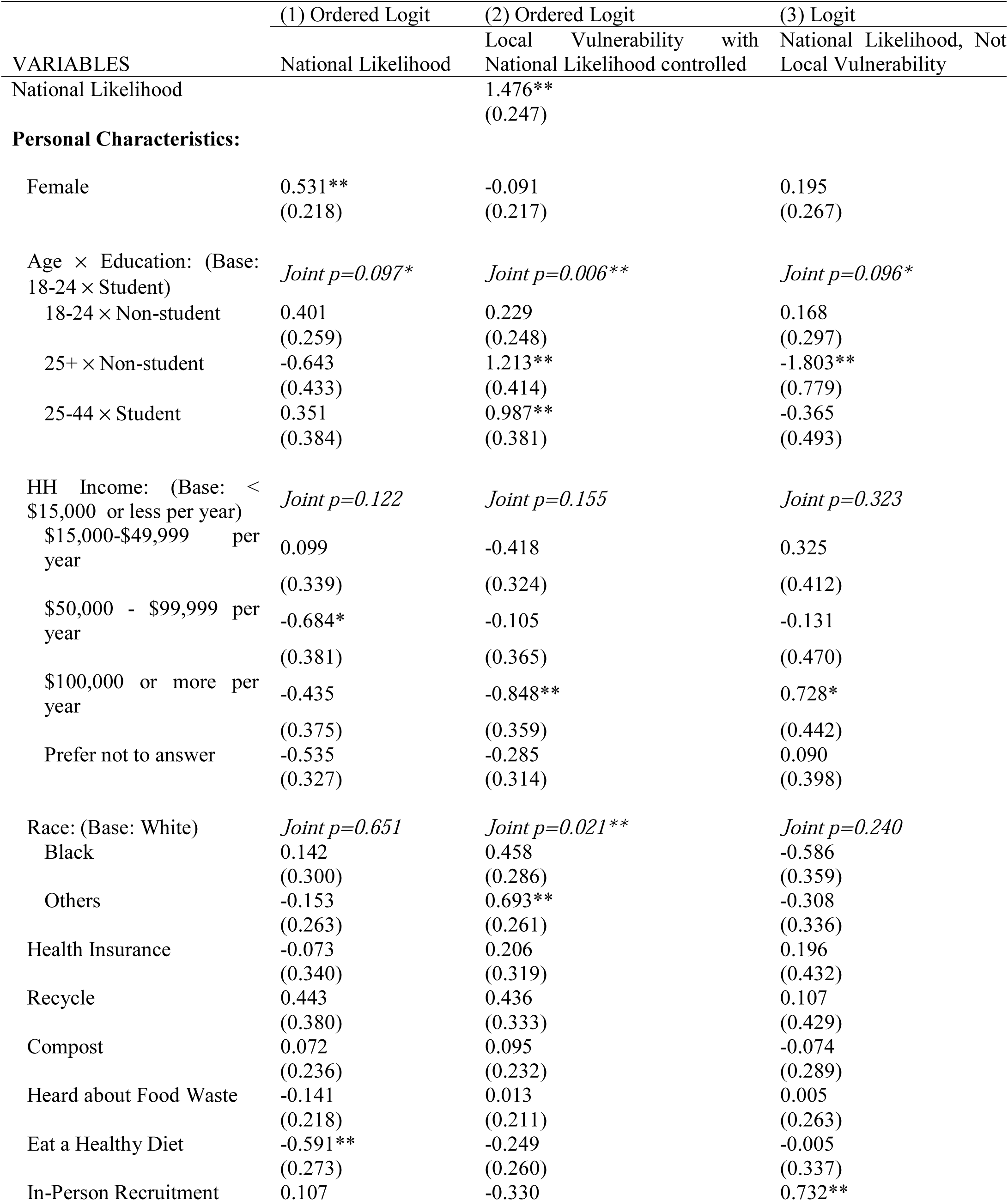
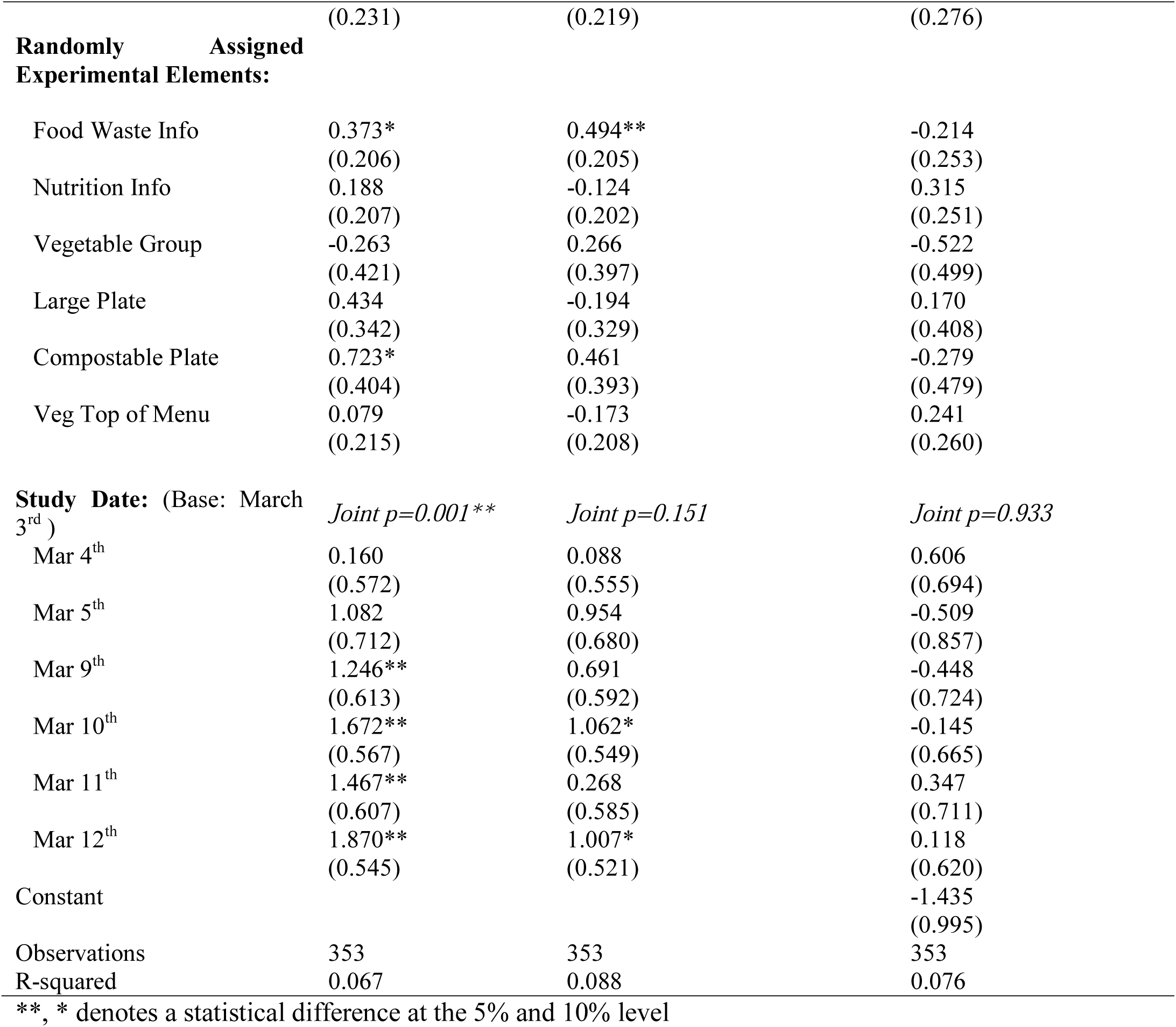
Regression models of COVID-19 question responses.

**Fig 3.**
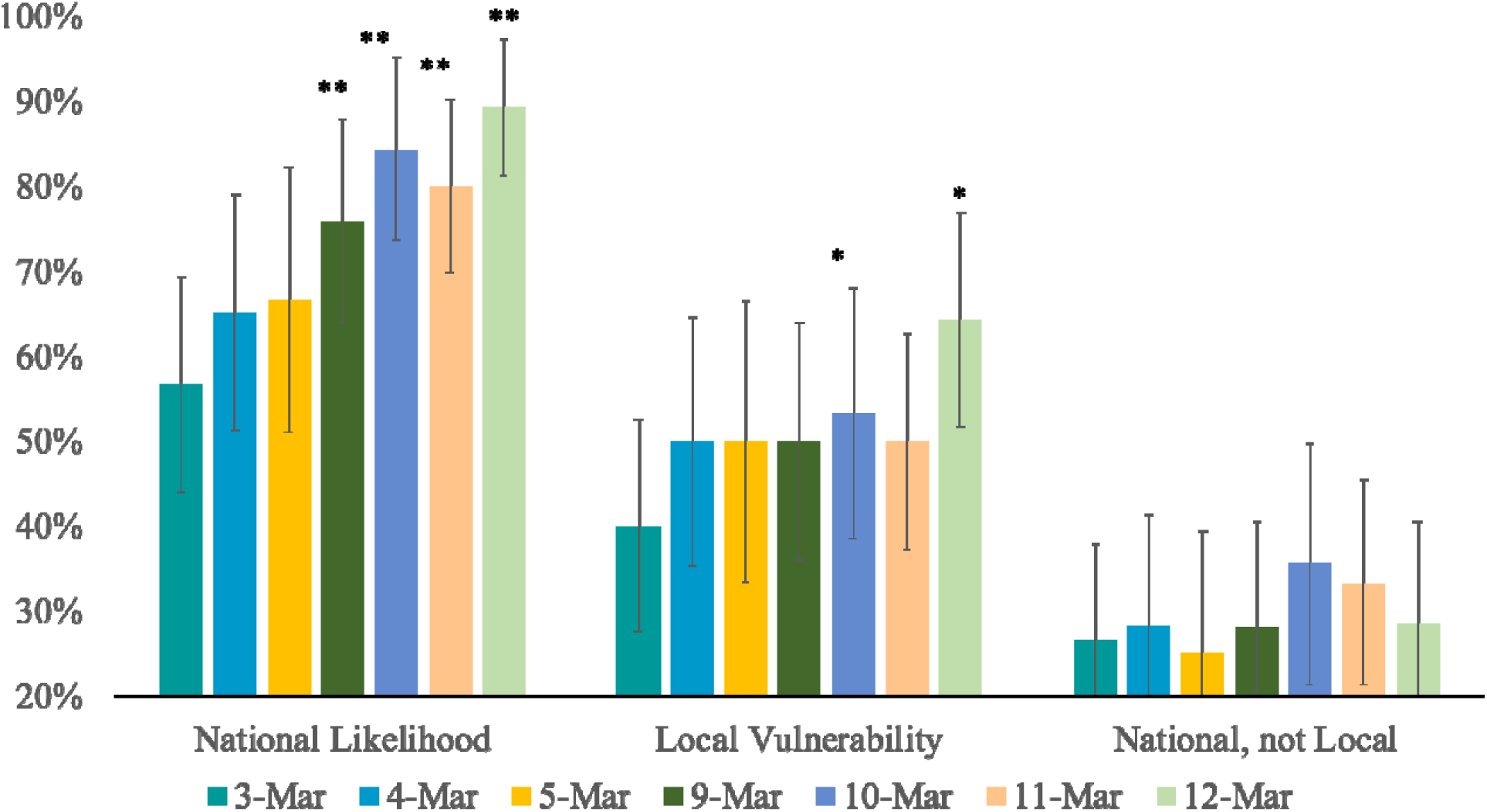
Daily Sample Means and 95% Confidence Intervals for Responses to COVID-19 Questions: (1) moderately or very likely that COVID-19 will cause a national public health crisis and (2) moderately or very concerned about contracting COVID-19 from attending campus events. The third group is the percent of participants who answered moderately/very likely and did not answer moderately/very concerned. 95% confidence interval bars do not control for covariates. **, * denotes a statistical difference of the value on this date from the value for the same variable on the first day of the study at the 5% and 10% level as determined by regression (Table 2) that controls for personal and experimental factors.

*Local Vulnerability* increased 10 percent points on March 4^th^, the day after Centers for Disease Control reported the potential public health threat posed by COVID-19 is very high to the United State and globally and expected more cases to be detected across the country, including more instances of person-to-person spread in more states [20]. Local Vulnerability remained relatively stable from March 4^th^ to March 11^th^, a timeframe during which COVID-19 cases increased more than 7-fold across the United States and participants’ perceived *National Likelihood* increased about 20 percent points. *Local Vulnerability* featured marginally significant increases on March 10, the day after the first presumptive positive case in Louisiana was announced [15], and on March 12, the day after the Governor of Louisiana declared a statewide public health emergency [17]. The percent of participants in the *National, not Local* response pattern (agreeing a national crisis was likely but not expressing concern about attending campus events) stayed relatively constant over the period and featured no significant differences from the first day of the study.

## Associations with COVID-19 Question Responses

Table 2 displays the estimated ordered logit results for *National Likelihood* and *Local Vulnerability* variables in their Likert scale form (1 = very unlikely/unconcerned, …., 5 = very likely/concerned) and binary logit model was estimated for the *National, not Local* variable.

The only personal characteristics that were significantly associated with *National Likelihood* were sex and diet. Men and those trying to eat a healthier diet provided lower likelihood ratings. The variables capturing the day of the study were jointly significant (*p* < 0.001) with each day during the second week significantly greater than the base (omitted) first day of the study. No randomly assigned experimental elements were significant, though several (food waste information, provision of compostable plates) were marginally significant.

*Local Vulnerability* was significantly and positively associated with *National Likelihood* such that a participant’s concern with contracting COVID-19 from attending campus events was greater as the individual participant’s likelihood of a national public health crisis increased. We modeled *National Likelihood* as an explanatory variable for *Local Vulnerability* because the *Local Vulnerability* question was asked immediately after the *National Likelihood* question. Personal characteristics that are positively associated with *Local Vulnerability* include being 25 years or older (regardless of student status) and identifying with a race other than white or black. Those in the highest income category ($100,000 or more) displayed significantly lower *Local Vulnerability* than those earning less than $15,000 per year. Participants that were randomly assigned the food waste information treatment (rather than the screen time information treatment) also reported significantly higher *Local Vulnerability*. The variables capturing the day of the study were not jointly significant and only March 10 and 12 were individually marginally significantly different from the omitted first day of the study.

Only two variables were significant in the regression model for *National, Not Local*. Older (≥ 25 years), non-students were less likely to feature this response pattern than younger students while those who attended the experiment in response to in-person flyers were more likely to feature this response pattern. No experimental treatments were significant, nor were there any significant differences by the day of the study.

## Classification Trees

Rather than making predictions based on *ceteris paribus* regression coefficients, the classification tree categorizes subjects based on splits from various predictor variables [21].

Figure 4 shows the pruned classification tree with a misclassification rate of 41% (i.e., 59% prediction accuracy). The first split is between those identifying as white versus other racial and ethnic identities, which represents the first determinant of expressing *Local Vulnerability*. Among those identifying as white and non-Hispanic, the study week in which they participated is the next branching variable, with those participating on March 10^th^ – 12^th^ being sorted based on whether they reported income less than $15,000 with the proportion in that lowest income group expressing *Local Vulnerability* being twice as high. For those attending March 9 or earlier, those randomly assigned to receive compostable plates were 72% more likely to express *Local Vulnerability* than those randomly assigned to other treatments.

**Fig 4.**
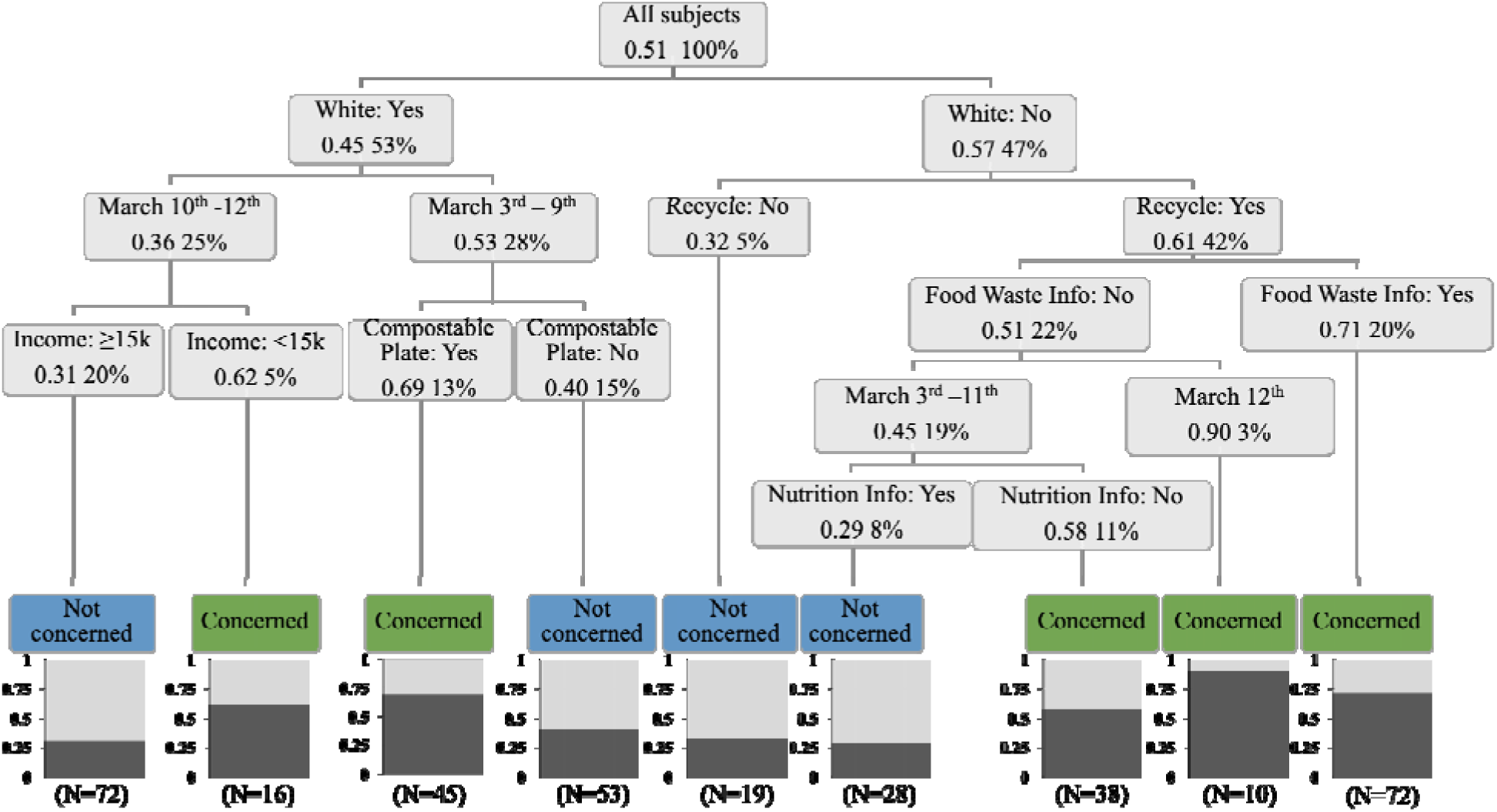
Classification Tree for *Local Vulnerability. Note*: Figures in each box are the proportion of participants in the regression tree branch that were very or moderately concerned about *Local Vulnerability* and the percent of participants falling into the branch. The bottom row features a bar graph of the proportion in that branch that were very or moderately concerned about *Local Vulnerability* and the number of participants in that branch. For example, 20% of participants (N=72) identified as white, responded on March 10^th^ -March 12^th^ and reported income ≥ $15,000, and the proportion of this group reported being very or moderately concerned about contracting COVID-19 from attending campus events was 0.31.

Among those identifying with groups other than white, non-Hispanic, self-reported recycling frequency was the next classification variable, with those reporting that they never recycle expressing less *Local Vulnerability* than most other branches. Those randomly receiving food waste information on any date were also in a branch with high *Local Vulnerability* as were those not receiving food waste information so long as it was on March 12^th^, the final day of the study.

## Discussion

As Poletti et al. [22] noted, the spread of epidemics can be dramatically delayed or mitigated if individual perception of the risk of the epidemic is sufficiently large and leads to reduced community contact. The authors emphasize that earlier public warnings about the epidemic can lead to dramatic reductions in peak prevalence and the final size of the infected population. Our analysis suggests that for LSU students, staff and faculty participating in a standard campus-based study unrelated to the topic of infectious disease, a majority of participants thought a national public health crisis from COVID-19 was moderately or very likely even on March 3, the first day of our data collection, a date upon which the number of cases nationally was reported to be 80 [14]. Nine days later, when the number of reported national cases exceeded 1600 [14], nearly 90% of participants displayed *National Likelihood*, with a significant jump in this perception compared to the first study day occurring at the beginning of the second week of the study (March 9).

Regression analysis reveals some personal characteristics significantly associated with *National Likelihood* that align with the previous literature, e.g., women perceive a national public health crisis as more likely than men [23-24]. Other significant associations have no precedent in the extant literature, e.g., participants who are trying to eat a healthier diet are significantly negatively associated with *National Likelihood*. This result may simply be spurious, or it may reflect a more nuanced relationship between dietary and health aspirations and national public health perceptions that we are unable to disentangle given the *post-hoc* nature of this analysis *vis a vis* the COVID-19 questions. For example, the result might reflect that those desiring to eat healthier have been exposed to wide-spread nutrition misinformation in the media, which has recently included unfounded claims that healthy eating or certain supplements reduce the likelihood of developing COVID-19. Indeed, the proliferation of dubious nutritional products has resulted in federal warnings to several companies promoting unproven nutrition-based remedies and preventatives for COVID-19 [25]. This might indicate reduced perceived risk of acquiring infectious disease among healthy eaters in our sample, based on these factors, although further investigation is required.

In Poletti et al.’s [22] model of epidemics, the spread is highly sensitive to the translation of risk perception to self-prophylaxis measures, such as social distancing, which slows community spread. While we did not elicit explicit measures of such behaviors, we did assess participants’ perceived vulnerability to contracting COVID-19 from attending campus events, (*Local Vulnerability*), which may signal a willingness to undertake social distancing and other beneficial behaviors and be a behavioral precursor. The first insight from observing the raw data plot in Figure 2 is that *Local Vulnerability* persistently lags *National Likelihood*, and does not significantly exceed the 50% mark until the last day of the study, which is the first day after the state of Louisiana had declared a public health emergency, but before LSU had cancelled classes or campus events.

Regression analysis confirms that national level perceptions are associated with perceived local vulnerability, as the *National Likelihood* variable in the *Local Vulnerability* regression features a large, significant and positive coefficient. In addition, participants 25 or older and those identifying with races other than white and black are more likely to express *Local Vulnerability*, while those in the highest income category expressed lower *Local Vulnerability* than those in the lowest income bracket. These results largely align with other findings from the literature. For example, Rhodes and Pivik [26] found drivers 25 and older perceived significantly higher risk from aggressive driving tactics than did younger drivers, while Lo [27] found that higher income respondents expressed less concern about environmental risks, which he hypothesized to stem from a heightened sense of material risk faced by those with lower incomes. Flynn, Slovic and Mertz [28] found respondents identifying as white, particularly white men, registered significantly lower environmental risk perceptions, hypothesizing that socio-political factors including power and status may influence risk perceptions.

Other characteristics typically identified in the literature (e.g., sex) are not significantly associated with expressed *Local Vulnerability*. Interestingly, participants randomly assigned an information treatment focused on the social and financial costs of food waste were significantly more likely to express *Local Vulnerability* and marginally higher on *National Likelihood*. While we cannot provide a definitive explanation of this relationship given *post-hoc* design constraints, we note the food waste information treatment was the only information treatment to emphasize national level and household level implications of individual behavior (e.g., food waste causing $161 billion of losses at the national level and $1500 of losses in an average household). Further, the classification tree finds that, similar to the randomly assigned information about food waste, participants who were randomly assigned compostable paper plates and the participant’s recycling habits also work as significant determinants of *Local Vulnerability*. One conjecture is that participants who link the implications of individual behaviors to issues of sustainability may reflect more critically on the implications of personal actions during a public health crisis, which could help increase compliance with social distancing and other preventative behaviors.

There is a persistent group consisting of about 30% of participants who, for the entire study period, including the final day, do not translate their perceived likelihood of a national public health crisis into personal vulnerability from attending campus events (*National, not Local*). These are likely a critical group in terms of modeling diffusion of COVID-19, as Poletti et al. [22] emphasize the role of translating perceived risk into preventative behaviors such as social distancing.

However, our analysis provides few insights into the characteristics associated with *National, not Local* group. Regression analysis finds few significant associations other than the fact that older non-students are less likely to feature this response pattern and that those who spontaneously attended the study in response to same-day receipt of flyers were more likely. The former suggests that younger people in academic settings may be diagnostic for predicting this response pattern while the latter may be suggestive that certain personality traits have predictive power.

This lack of insight into the *National, not Local* group is likely due to the post-hoc nature of the analysis, one of several study limitations. Specifically, the study was originally designed to investigate a topic other than COVID-19 perceptions, hence logical experimental treatments and additional questions about personal perceptions and behaviors relevant to understanding and predicting the spread of COVID-19 were not included and the questions that were posed were not motivated by theory. Another study limitation is that the sample is drawn from a single academic institution, limiting the representativeness of the data geographically, demographically, and socioeconomically. Finally, the data were acquired prior to the declaration of a national emergency, and we would expect further evolution in how people in this location might respond to these questions in the face of more dire national promulgations concerning the pandemic.

## Conclusions

By integrating questions focused on COVID-19 into an ongoing in-person experiment during the two weeks prior to the major disruption in public activities in Louisiana and much of the country, we provide some insights into how participants drawn from one community in Louisiana were perceiving the national and local implications of the public health crisis that was unfolding during the study period. Understanding perceptions related to risk can help to tailor national or local responses to curb transmission of infectious disease.

We find that perceptions during this critical time increased steadily and rapidly such that nearly 90 percent of participants agreed that it was likely that COVID-19 would become a national public health crisis by the final day of our study, which corresponded with the day that Louisiana declared a public health emergency. However, participants’ views of their personal vulnerability to contracting the virus from attending local events increased more slowly and, only on the day after Louisiana’s emergency declaration, did a majority of participants agree that public event attendance increased their odds of contracting the virus.

While some characteristics that were significantly associated with a lower perceived local vulnerability to contracting COVID-19 have precedent from previous risk perception research (e.g., younger than 25, white, higher incomes), others are novel and suggest the need for more investigation. For example, our finding of significantly lower perceived local vulnerability among participants expressing a strong interest in eating healthier may support aggressive information and enforcement campaigns against dietary schemes promoting themselves as COVID-19 preventatives or remedies or broadly touting certain foods as immunity-boosting [25]. Also, our finding that participants who were randomly assigned an information treatment that emphasized the national implications of food waste expressed significantly higher perceptions of local vulnerability may suggest that information campaigns emphasizing the national implications of individual behaviors could help increase compliance with social distancing and other preventative behaviors.

Throughout the study period, including the day after the emergency declaration, and about 30% of participants did not convert national perceptions of a likely public health crisis into perceived vulnerability from local event attendance. This could be a key group to target as localities and states implement social distancing policies and procedures. The significant characteristics associated with this group are limited, but do include age, with students less than 25 years of age more likely to fall into this group than older, non-students. This provides evidence to support strategies that tailor communications efforts to younger cohorts that encourage social distancing and other prevention behaviors (e.g., WHO 2020 [29]).

## Data Availability

Is available in the supplemental materials that will accompany any published article.

## Acknowledgements

The authors thank Adriana Alfaro, Ana Lucia Gutierrez, Erika Largacha, Estela Barahona, De’Jerra Bryant, Nila Pradhananga, and Runlan Cai for excellent research assistance. All remaining errors are those of the authors.

## Supporting Information

**S1 appendix. Exit survey**.

**S2 appendix. Information and experimental treatments**.

